# Training deep learning algorithms with weakly labeled pneumonia chest X-ray data for COVID-19 detection

**DOI:** 10.1101/2020.05.04.20090803

**Authors:** Sivaramakrishnan Rajaraman, Sameer Antani

**Affiliations:** Lister Hill National Center for Biomedical Communications, National Library of Medicine, 8600 Rockville Pike, Bethesda, MD 20894, USA

**Keywords:** augmentation, chest-X-rays, convolutional neural network, COVID-19, deep learning, pneumonia, localization

## Abstract

The novel Severe Acute Respiratory Syndrome Coronavirus 2 (SARS-CoV-2) has caused a pandemic resulting in over 2.7 million infected individuals and over 190,000 deaths and growing. Respiratory disorders in COVID-19 caused by the virus commonly present as viral pneumonia-like opacities in chest X-ray images which are used as an adjunct to the reverse transcription-polymerase chain reaction test for confirmation and evaluating disease progression. The surge places high demand on medical services including radiology expertise. However, there is a dearth of sufficient training data for developing image-based automated decision support tools to alleviate radiological burden. We address this insufficiency by expanding training data distribution through use of weakly-labeled images pooled from publicly available CXR collections showing pneumonia-related opacities. We use the images in a stage-wise, strategic approach and train convolutional neural network-based algorithms to detect COVID-19 infections in CXRs. It is observed that weakly-labeled data augmentation improves performance with the baseline test data compared to non-augmented training by expanding the learned feature space to encompass variability in the unseen test distribution to enhance inter-class discrimination, reduce intra-class similarity and generalization error. Augmentation with COVID-19 CXRs from individual collections significantly improves performance compared to baseline non-augmented training and weakly-labeled augmentation toward detecting COVID-19 like viral pneumonia in the publicly available COVID-19 CXR collections. This underscores the fact that COVID-19 CXRs have a distinct pattern and hence distribution, unlike non-COVID-19 viral pneumonia and other infectious agents.

## 1. Introduction

The novel Coronavirus disease 2019 (COVID-19) is caused by a strain of coronavirus called the Severe Acute Respiratory Syndrome Coronavirus 2 (SARS-CoV-2) that originated in Wuhan in the Hubei province in China. On March 11, 2020, the World Health Organization (WHO) declared the disease as a pandemic [1], and as of this writing (in late April 2020), there are more than 2.7 million globally confirmed cases with over 190,000 reported deaths with unabated growth. The disease is detected using the reverse transcription-polymerase chain reaction (RT-PCR) tests that are shown to exhibit high specificity but variable sensitivity in detecting the presence of the disease [2]. However, these test kits are in limited supply in some geographical regions, particularly third-world countries [3]. The turnaround time is reported to be 24 hours in major cities and even greater in rural regions. This necessitates the need to explore other options to identify the disease and facilitate swift referrals for the COVID-19 affected patient population in need of urgent medical care.

A study of literature shows that viral pneumonia is commonly found to affect the lungs with the progression of COVID-19 disease, often manifesting as ground-glass opacities (GGO), with peripheral, bilateral, and predominant basal distribution in the lungs, preventing oxygen entry, thereby causing breathing difficulties along with hyperthermia [2]. These patterns are visually similar to, yet distinct from those caused by non-COVID-19-related viral pneumonia and those caused by other bacterial and fungal pathogens [2]. Also, current literature studies reveal that it is difficult to distinguish viral pneumonia from others caused by bacterial and fungal pathogens [4]. Fig. 1 shows instances of CXRs with clear lungs, showing bacterial pneumonia, and COVID-19-related pneumonia, respectively.

**Figure 1.**
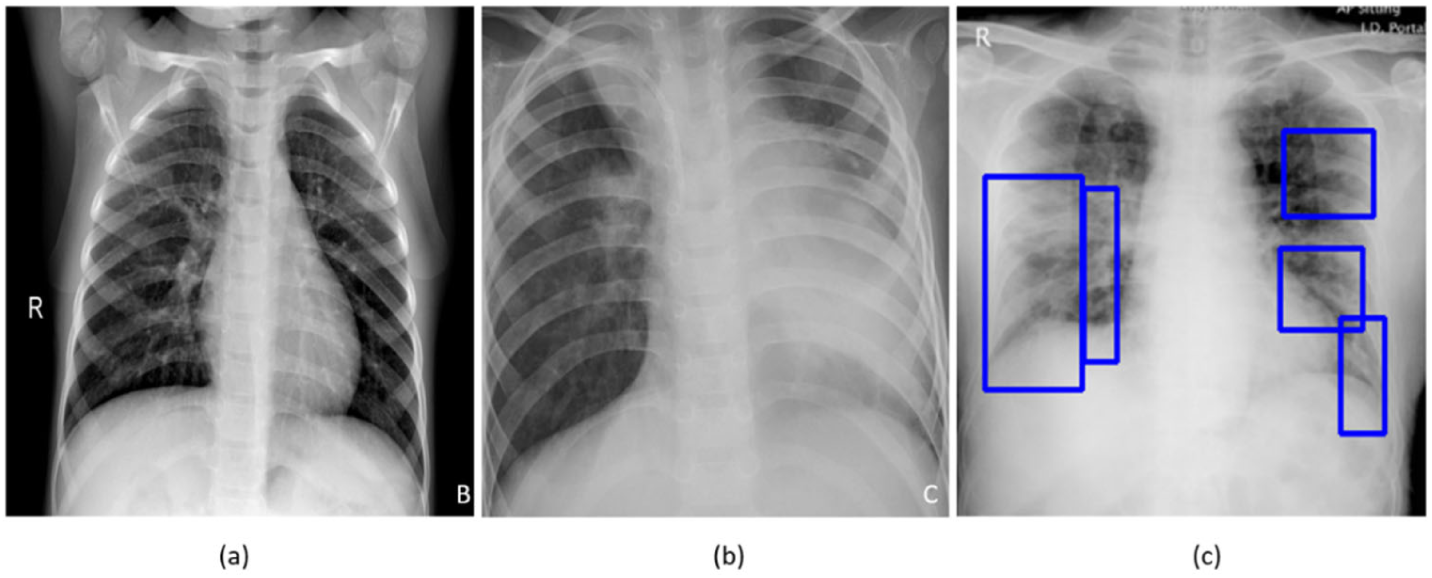
CXRs showing (**a**) Clear lungs; (**b**) Bacterial pneumonia infections manifesting as consolidations in the right upper lobe and retro-cardiac left lower lobe; (**c**) COVID-19 pneumonia infection showing bilateral manifestations.

While not recommended as a primary diagnostic tool due to risk of increased transmission, chest radiography and computed tomography (CT) scans are used to screen/confirm respiratory damage in COVID-19 disease and evaluate its progression [3]. CT scans are reported to be less specific than RT-PCR but highly sensitive in detecting COVID-19 and can play a pivotal role in disease diagnosis/treatment [3]. However, the American College of Radiology has recommended against use of CT scans as a first-line test^1^. Additional considerations of increased risk of transmission, access, and cost also contribute to the recommendation. When radiological imaging is considered necessary, portable chest X-rays (CXRs) are considered a good and viable alternative [2]. However, in a pandemic situation, assessment of the images places a huge burden on radiological expertise, which is often lacking in regions with limited resources. Automated decision-making tools could be valuable in alleviating some of this burden, and also as a research tool for quantifying disease progression.

A study of literature shows that automated computer-aided diagnostic (CADx) tools built with data-driven deep learning (DL) algorithms using convolutional neural networks (CNN) have shown promise in detecting, classifying, and quantifying COVID-19-related disease patterns using CXRs and CT scans [5, 6] and can serve as a triage under resource-constrained settings thereby facilitating swift referrals that need urgent patient care. These tools combine elements of radiology and computer vision to learn the hierarchical feature representations from medical images to identify typical disease manifestations and localize suspicious regions of interest (ROI).

It is customary to train and test a DL model with the data coming from the same target distribution to offer probabilistic predictions toward categorizing the medical images to their respective categories. Often this idealized target is not possible due to limited data availability, or weak labels. In the present situation, despite a large number of cases worldwide, we have very limited COVID-19 CXR image data that is publicly available to train DL models where the goal is to recognize CXR images showing COVID-19-related viral pneumonia from those caused by other non-COVID-19 viral, bacterial and other pathogens. Acquiring such data remains a goal for medical societies such as the Radiological Society of North America (RSNA)^2^ and Imaging COVID-19 AI Initiative in Europe.^3^ Large number of training data enable a diversified feature space across categories that help enhance inter-class variance leading to better DL performance. The absence of such data leads to model overfitting and poor generalization to unseen real-world data. Under these circumstances, data augmentation has been proven to be effective in training discriminative DL models [7]. There are several data augmentation methods discussed in the literature for improving performance in natural computer vision tasks. These include traditional augmentation techniques like flipping, rotations, color jittering, random cropping, and elastic distortions and generative adversarial networks (GAN) based synthetic data generation [8].

Unlike natural images, such as those found in ImageNet [9], medical images tend to have different visual characteristics exhibiting high inter-class similarities and highly localized ROI. Under these circumstances, traditional augmentation methods that introduce simple pixel-wise image modifications are shown to be less effective [10]. On the other hand, GAN-based DL models that are used for synthetic data generation are computationally complex and the jury is still out on the anatomical and pathological validity of synthesized images. These networks are hard to train due to the problem of Nash equilibria, defined as the zero-sum game between the generator and the discriminator networks where they contest with each other in improving performance [11]. Further, these networks are shown to be sensitive to the selection of architecture and hyperparameters and often get into mode collapse, resulting in degraded performance [11]. In general, there is a great opportunity for research in developing effective data augmentation strategies for medical visual recognition tasks. Goals for such medical data augmentation techniques include reducing overfitting and regularization errors in a data-scarce situation. The urgency offered by the pandemic has led to the motivation behind this study.

In this work, we use weakly-labeled CXR images that are pooled from publicly available collections showing pneumonia-related opacities to augment training data toward improving inter-class variance. The goal is to improve COVID-19 detection in CXRs, with the baseline being the training data without augmentation.

## 2. Materials and Methods

### 2.1. Data and Workflow

This retrospective analysis is performed using four publicly available CXR collections:

A. Pediatric CXR dataset [4]: A set of 5,232 anterior-posterior (AP) projection CXR images of children of 1 to 5 years of age acquired as part of routine clinical care at the Guangzhou Children’s Medical Center in China. The set contains 1583 normal, 2780 bacterial pneumonia, and 1493 CXRs showing non-COVID-19 viral pneumonia, respectively.
B. RSNA CXR dataset [12]: The RSNA, Society of Thoracic Radiology (STR), and the National Institutes of Health (NIH) jointly organized the Kaggle pneumonia detection challenge to develop image analysis and machine learning algorithms to automatically categorize the CXRs as showing normal, non-pneumonia-related or pneumonia-related opacities. The publicly available data is a curated subset of 26,684 AP and posterior-anterior (PA) CXRs showing normal and abnormal radiographic patterns, taken from the NIH CXR-14 dataset [13]. It includes 6012 CXRs showing pneumonia-related opacities with ground truth (GT) bounding box annotations for these on 1,241 CXRs.
C. CheXpert CXR dataset [14]: A subset of 4683 CXRs showing pneumonia-related opacities selected from a collection of 223,648 CXRs in frontal and lateral projections, collected from 65,240 patients at Stanford Hospital, California, and labeled for 14 thoracic diseases by extracting the labels from radiological texts using an automated natural language processing (NLP)-based labeler, conforming to the glossary of the Fleischner Society.
D. NIH CXR-14 dataset [13]: A subset of 307 CXRs showing pneumonia-related opacities selected from a collection of 112,120 CXRs in frontal projection, collected from 30,805 patients. Images are labeled with 14 thoracic disease labels extracted automatically from radiological reports using an NLP-based labeler.
E. Twitter COVID-19 CXR dataset: A collection of 135 CXRs showing COVID-19-related viral pneumonia, collected from SARS-CoV-2 positive subjects has been made available by a cardiothoracic radiologist from Spain via Twitter. (https://twitter.com/ChestImaging) The images are made available in JFIF format at approximately 2K×2K resolution.
F. Montreal COVID-19 CXR dataset: As of April 14, 2020, a collection of 179 SARS-CoV-2 positive CXRs and others showing non-COVID-19 viral disease manifestations has been made publicly available by the authors of [15] in their GitHub repository. The CXRs are made available in AP and PA projections.

Table 1 shows the distribution of data extracted from the datasets identified above and used for the different stages of learning performed in this study. The numerator and denominator show the number of train and test data used in models’ training and evaluations. The GT disease bounding box annotations for a sample of the test data, containing 27 CXRs collectively from the Twitter COVID-19 and Montreal COVID-19 CXR collections is set by the verification of publicly identified cases from an expert radiologist who annotated the sample test collection.

**Table 1.** Dataset characteristics. Numerator and denominator denote the number of train and test data respectively (UP=Pneumonia of unknown type, BP= Bacterial (proven) pneumonia, VP= non-COVID-19 viral (proven) pneumonia, CP = COVID-19 pneumonia).

| Dataset | UP | BP | VP | CP |
| --- | --- | --- | --- | --- |
| A | - | 2538/242 | 1345/148 | - |
| B | -/6012 | - | - | - |
| C | -/4683 | - | - | - |
| D | -/307 | - | - | - |
| E | - | - | - | -/135 |
| F | - | - | - | -/179 |

Broadly, our workflow consists of the following steps: First, we preprocess the images to make them suitable for use in DL. Then, we evaluate the performance of a custom CNN and a selection of pre-trained CNN models for binary categorization of the publicly available pediatric CXR collection showing bacterial or viral pneumonia. The trained model is further used to categorize the publicly available COVID-19 CXR collections as showing viral pneumonia. Next, we use the trained model to weakly label the CXRs in the publicly available CXR collections with pneumonia-related opacities as showing bacterial or viral pneumonia. The baseline training data is augmented with these weakly labeled CXRs to improve detection performance with the baseline hold-out test data and the COVID-19 CXR collections. We also augment the baseline training with COVID-19 CXRs from one of the two different collections to evaluate for an improvement in performance in detecting CXRs showing COVID-19 viral pneumonia from the other collection. This data augmentation strategy recognizes the biological similarity in viral pneumonia and radiological manifestation due to COVID-19 caused respiratory disease. It also takes advantage of dissimilarity to bacterial pneumonia-related opacities. Finally, the strategy reduces the intra-class similarity and enhances inter-class discrimination in the strategic ordering of the coarsely labeled data. We have already shown in our other work that iteratively pruned deep learning ensembles produce impressive results with this data [6]. In this work, we show that it is also possible to obtain very good results using a biologically sensitive and discriminative training data augmentation strategy.

### 2.2. Lung ROI Segmentation and Preprocessing

It is important to add controls during training data-driven DL methods for disease screening/diagnosis. Learning irrelevant feature representations could adversely impact clinical decision making. To assist the DL model to focus on pulmonary abnormalities, we used a dilated dropout-U-Net [16] to segment the lung ROI from the background. Dilated convolutions are shown to improve performance [17] with exponential receptive field expansion while preserving spatial resolution with no added computational complexity. A Gaussian dropout with an empirically determined value of 0.2 is used after the convolutional layers in the network encoder to avoid overfitting and improve generalization. A publicly available collection of CXRs and their associated lung masks [18] is used to train the dilated dropout-U-Net model to generate lung masks of 224×224 pixel resolution. Callbacks are used to store the best model weights after each epoch. The generated masks are superimposed on the original CXRs to delineate the lung boundaries, crop them to the size of a bounding box, and re-scale them to 224×224 pixel resolution to reduce computational complexity. Fig. 2 shows the segmentation steps performed in this study.

**Figure 2.**
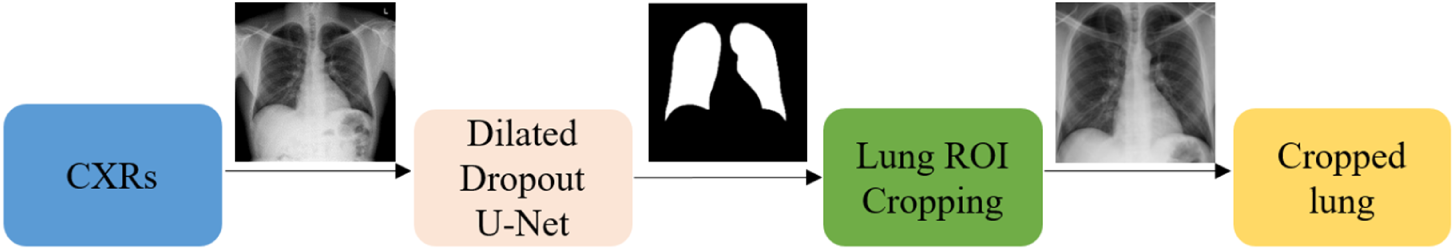
The segmentation approach showing dilated dropout U-Net based mask generation and Lung ROI cropping.

Additional preprocessing steps performed are as follows: i) CXRs are thresholded at to remove very bright pixels to remove text annotations (empirically determined to be in the range [235 255]) that might be present in the cropped images. Missing pixels are in-painted using the surrounding pixel values. ii) Images are normalized to make the pixel values lie in the range [0, 1]. iii) CXR images are median filtered to remove noise and preserve edges. iv) Image pixel values are centered and standardized to reduce computational complexity. Next, the cropped CXRs are used to train and evaluate a custom CNN and a selection of pretrained models at different learning stages performed in this study.

### 2.3. Models and Computational Resources

The performance of a custom CNN model whose design is inspired by wide residual network (WRN) architecture proposed in [19] and a selection of ImageNet pretrained CNN models is evaluated during different stages of learning performed in this study. The benefit of using a WRN compared to the traditional residual networks (ResNets) [20] is that it is shallower resulting in shorter training times while producing similar or improved accuracy. In this study, we used a WRN based custom CNN architecture with dropouts used in every residual block. After pilot empirical evaluations, we used a network depth of 28, a width of 10, and a dropout ratio of 0.3 for the custom WRN used in this study.

We evaluated the performance of the following pretrained CNN models, viz., a) VGG-16 [21], b) Inception-V3 [22], c) Xception [23], d) DenseNet-121 [24], and e) NasNet-mobile [25]. The pretrained CNNs are instantiated with their ImageNet [9] pretrained weights and truncated at their fully-connected layers. The output feature maps are global average pooled and fed to a final dense layer with Softmax activations to output the prediction probabilities.

The following hyperparameters of the custom WRN and pretrained CNNs are optimized through a randomized grid search method: i) momentum, ii) L2-weight decay, and iii) initial learning rate of the Stochastic Gradient Descent (SGD) optimizer. We initialized the search ranges to [0.80 0.99], [1e-8 1e-2], and [1e-7 1e-3] and for the learning momentum, L2-weight decay, and initial learning rate, respectively. The custom WRN is initialized with random weights and the pretrained models are fine-tuned end-to-end with smaller weight updates to make them data-specific and classify the CXRs to their respective categories. Callbacks are used to monitor model performance and store the best model weights for further analysis.

The performance of the custom WRN and the pretrained CNN models are evaluated in terms of i) accuracy, ii) area under the (receiver operating characteristic -- ROC) curve (AUC), ii) sensitivity or recall, iv) specificity, v) precision, vi) F-score, and vii) Mathews correlation coefficient (MCC). The models are trained and evaluated on a Windows System with Intel Xeon CPU 3.80 GHz with 32 GB RAM and NVIDIA GeForce GTX 1070 GPU. We used Keras 2.2.4 API version with Tensorflow backend and CUDA/CUDNN dependencies.

### 2.4. Weakly-labeled Data Augmentation

We train the custom WRN and the pretrained models on the pediatric CXR collection [4] and evaluated them on the ability to categorize hold-out test data into bacterial and viral pneumonia categories. This start stems from following the literature which reveals that CXRs showing COVID-19 viral pneumonia manifestations are visually similar to, yet distinct from those caused by bacterial, fungal, and other non-COVID-19-related viral pneumonia [2]. We use the best performing baseline model to evaluate its performance in categorizing the CXRs from Twitter COVID-19 and Montreal COVID-19 collections as belonging to the viral pneumonia category.

We also evaluated the performance of the best performing baseline model in weakly categorizing the CXRs showing pneumonia-related opacities from RSNA, CheXpert, and NIH CXR collections as belonging to the bacterial or viral pneumonia categories. These weakly classified CXRs are used to augment the baseline training data. The idea behind this augmentation is to expand the training data feature space: i) to make the training distribution encompass the variability in the test distribution, enhance inter-class discrimination, and reduce intra-class similarity; and, ii) to decrease the generalization error by training with samples from a diversified distribution. The model is trained with different combinations of the augmented training data and evaluated for an improvement in performance as compared to the non-augmented baseline in classifying: i) the baseline hold-out pediatric CXR test data to bacterial or viral pneumonia categories; and, ii) Twitter COVID-19 and Montreal COVID-19 CXR collections as belonging to the viral pneumonia category. The baseline training data is also augmented with the CXRs showing COVID-19 viral pneumonia from one of the two different COVID-19 CXR collections used in this study to evaluate for performance improvement with the other collection. This is done to evaluate if the COVID-19 viral pneumonia patterns are very distinct and unique that can only improve performance toward COVID-19 detection as compared to that with weakly-labeled data augmentation and non-augmented training.

### 2.5. Salient ROI Localization

Visualization helps in interpreting the model predictions and identify the salient ROI involved in decision-making. In this study, the learned behavior of the best performing baseline model in categorizing the CXRs to the bacterial and viral pneumonia classes is visualized through gradient-weighted class activation maps (Grad-CAM) [26]. Grad-CAM is a gradient-based visualization method where the gradients for a given class are computed concerning the features extracted from the deepest convolutional layer in a trained model and are fed to a global average pooling layer to obtain the weights of importance involved in decision-making. This results in a two-dimensional heat map which is a weighted combination of the feature maps involved in categorizing the image to its respective class.

## 3. Results and Discussion

Optimal hyperparameters values obtained using a randomized grid search for the custom WRN and pretrained CNNs that are trained and evaluated on the pediatric CXR collection to classify them at the patient level into showing bacterial or viral pneumonia are shown in Table 2. For model validation, we allocated 20% of the training data which was randomly selected. The performance achieved by the models is shown in Table 3.

**Table 2.** Optimal values for the hyperparameters for the custom WRN and pretrained CNNs obtained through randomized grid search M: Momentum, ILR: Initial learning rate, and L2: L2-weight decay).

| Models | Optimal values |  |  |
| --- | --- | --- | --- |
|  | M | ILR | L2 |
| Custom | 0.90 | 1e-3 | 1e-5 |
| Pretrained | 0.95 | 1e-3 | 1e-6 |

**Table 3.** Performance achieved by the custom WRN and pretrained CNNs in classifying the pediatric CXR dataset into bacterial and viral categories. Here Acc.: Accuracy, Sens.: Sensitivity, Prec.: Precision, F: F-score, and MCC: Matthews Correlation Coefficient). Here bold values indicate superior performance.

| Models | Acc. | AUC | Sens. | Spec. | Prec. | F | MCC |
| --- | --- | --- | --- | --- | --- | --- | --- |
| Custom WRN | 0.8974 | 0.9534 | 0.9381 | 0.8311 | 0.9008 | 0.9191 | 0.7806 |
| VGG-16 | **0.9308** | **0.9565** | 0.9711 | 0.8649 | 0.9216 | **0.9457** | **0.8527** |
| Inception-V3 | 0.9103 | 0.937 | 0.9587 | 0.8311 | 0.9028 | 0.9299 | 0.8085 |
| Xception | 0.9282 | 0.954 | 0.9546 | **0.8852** | **0.9315** | 0.9429 | 0.8469 |
| DenseNet-121 | 0.9026 | 0.9408 | 0.967 | 0.7973 | 0.8864 | 0.925 | 0.7931 |
| NASNet-mobile | 0.9282 | 0.9479 | **0.9753** | 0.8514 | 0.9148 | 0.944 | 0.8477 |

It can be observed that the VGG-16 model demonstrates superior performance in terms of accuracy and AUC with the hold-out test data. Xception model gives higher precision and specificity than the other models. However, considering the F-score and MCC that give a balanced precision and sensitivity measure, the VGG-16 model outperformed the others in classifying the pediatric CXRs as showing bacterial or viral pneumonia. The performance excellence of the VGG-16 model is attributed to the fact that the architecture depth of the model is optimal to learn from the data used in this study and extract diversified features to categorize the CXRs to their respective categories. Deeper models like DenseNet-121 showed performance degradation as they suffered from overfitting issues and are not able to effectively model the variations across the categories. In this regard, we select the best performing VGG-16 model for further analysis on the Twitter COVID-19 and Montreal COVID-19 CXR collections as showing viral pneumonia.

In this part of the study, we establish a baseline using the learned representations for the viral pneumonia category from the pediatric CXR collection for identifying COVID-19 viral pneumonia-related manifestations in the aforementioned COVID-19 CXR collections. As mentioned before, this is based on the knowledge that COVID-19 is a kind of viral pneumonia, but while being similar is different in some respects [2]. The baseline performance achieved is shown in Table 4. Fig. 3 shows the confusion matrix obtained toward classifying Twitter and Montreal COVID-19 CXR collections as showing viral pneumonia using baseline VGG-16 model trained to separate bacterial from viral pneumonia in CXR images.

**Table 4.** Performance metrics achieved in classifying the Twitter and Montreal COVID-19 CXR collections as showing viral pneumonia using baseline VGG-16 model trained to separate bacterial from viral pneumonia in CXR images.

| Model | Accuracy |  |
| --- | --- | --- |
|  | Twitter-COVID-19 | Montreal-COVID-19 |
| VGG-16 | 0.2885 | 0.5028 |

**Figure 3.**
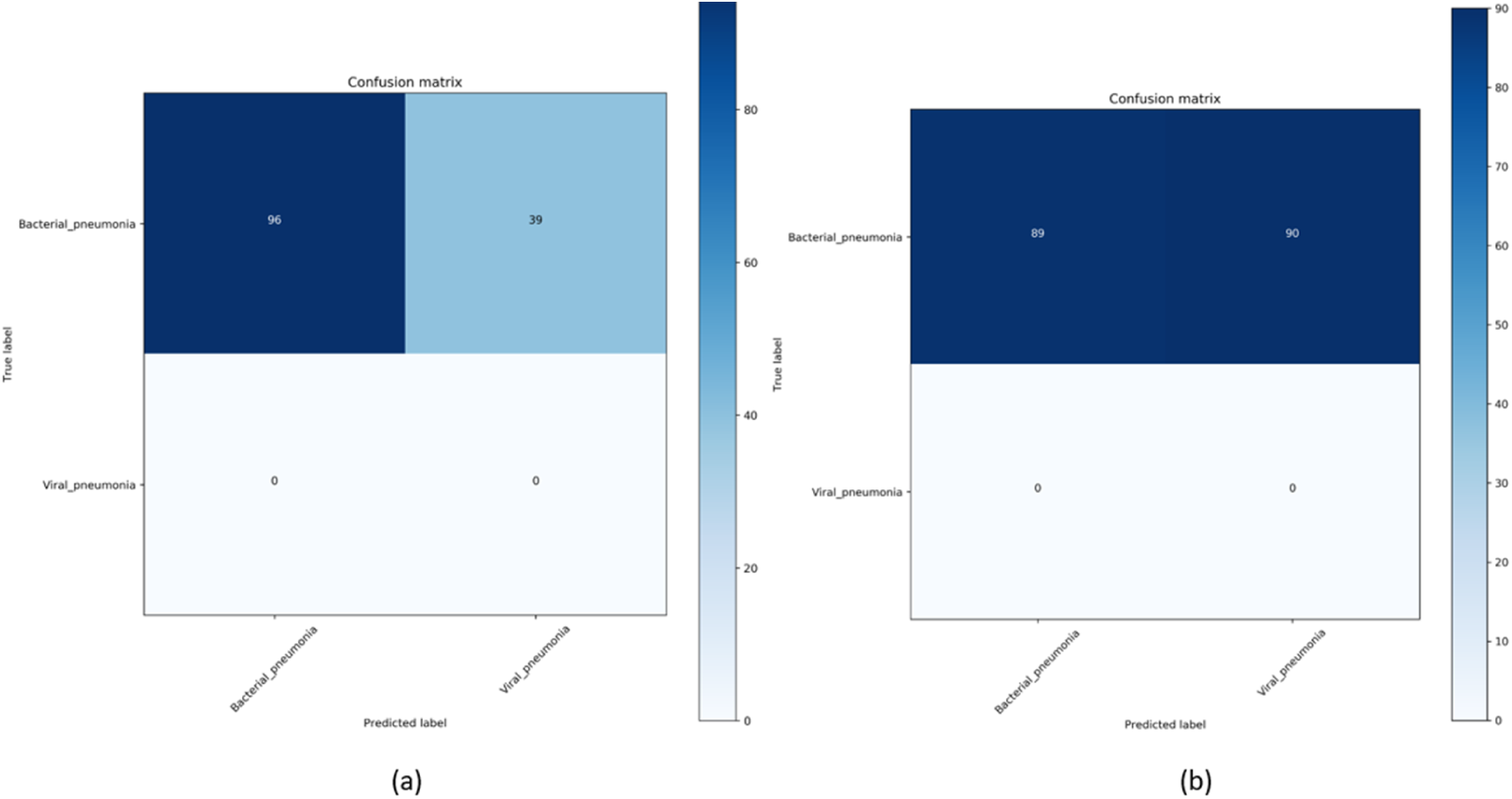
Confusion matrix obtained toward classifying (a) Twitter and (b) Montreal COVID-19 CXR collections as showing viral pneumonia using baseline VGG-16 model.

As observed in Table 4 and Fig. 3, the results obtained with the baseline VGG-16 model trained on the pediatric CXR collection to learn the representations of bacterial and viral pneumonia didn’t deliver superior performance in detecting COVID-19 related viral pneumonia manifestations in the Twitter and Montreal COVID-19 CXR collections. We attribute this to limited variance in the training distribution and hence a narrow feature space to learn related patterns. The model fails to appropriately classify the Twitter and Montreal COVID-19 CXR collections as belonging to the viral pneumonia class.

The learned behavior of the baseline trained VGG-16 model with the pediatric CXR collection is interpreted through Grad-CAM visualizations and is shown in Fig. 4. The gradients for the bacterial and viral pneumonia classes that are flowing into the deepest convolutional layer of the trained model are used to interpret the neurons involved in decision-making. The heat maps obtained as a result of weighing these feature maps are superimposed on the original CXRs to identify the salient ROI involved in categorizing the CXRs to their respective classes. It is observed that the model is correctly focusing on the salient ROI for the test data coming from the same training distribution that helps to categorize them into bacterial and viral pneumonia classes. However, the salient ROI involved in categorizing a test image from the Montreal COVID-19 CXR collection that comes from a different distribution compared to the baseline training data didn’t properly overlap with the GT annotations. This leads to the inference that the model is not properly trained to identify the disease manifestations in the unseen test data that has similar, yet distinct visual representations as to the baseline training data.

**Figure 4.**
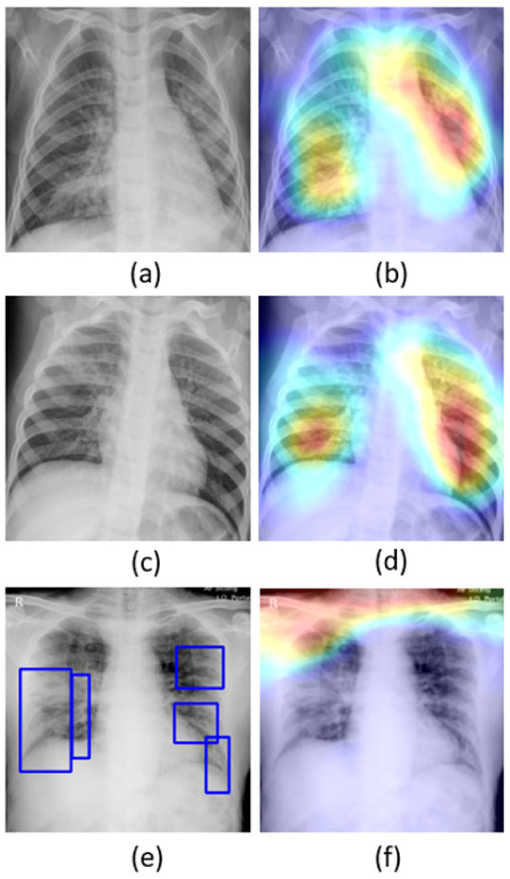
Original CXRs and their salient ROI visualization: (**a**) and (**b**) shows a CXR with bilateral bacterial pneumonia and the corresponding Grad-CAM visualization; (**c**) and (**d**) shows a CXR with viral pneumonia manifestations and the corresponding salient ROI visualization; (**e**) and (**f**) shows a sample CXR from the test set of Montreal COVID-19 CXR collection with GT annotations and the corresponding salient ROI visualization.

With data-driven DL methods, the training data may contain samples that do not contribute to decision-making. Modifying the training distribution could provide an active solution to improve performance with similar and/or different test distribution. In response, our approach is to expand the training data feature space to create a diversified distribution that could help learn and improve the performance with the baseline test data coming from the same distribution as the training data and/or with other test data coming from a different distribution. In this study, we propose to expand the training data feature space by augmenting them with weakly classified CXR images. For this, the trained baseline VGG-16 model is used to weakly classify the CXR images from NIH, RSNA, and CheXpert collections showing pneumonia-related opacities as showing bacterial or viral pneumonia. The weakly labeled images are further stored to augment the baseline training data to improve performance in categorizing the test data from pediatric, Twitter COVID-19, and Montreal COVID-19 CXR collections. We also augmented the baseline with the COVID-19 CXR collections to study their effect on improving performance with the baseline test data. The performance metrics achieved with the baseline test data using different combinations of the augmented training data is shown in Table 5.

**Table 5.** Performance metrics achieved with the different combinations of the augmented training data toward classifying the baseline test data into bacterial and viral pneumonia categories. Bold values indicate superior performance.

| Dataset | Acc. | AUC | Sens. | Spec. | Prec. | F | MCC |
| --- | --- | --- | --- | --- | --- | --- | --- |
| Baseline | 0.9308 | 0.9565 | 0.9711 | 0.8649 | 0.9216 | 0.9457 | 0.8527 |
| Data augmentation with weakly labeled images |  |  |  |  |  |  |  |
| Baseline + Montreal | 0.9179 | 0.9479 | 0.9794 | 0.8176 | 0.8978 | 0.9368 | 0.827 |
| Baseline + Twitter | 0.9308 | 0.9577 | 0.9835 | 0.8446 | 0.9119 | 0.9464 | 0.8541 |
| Baseline + NIH | 0.9179 | 0.9600 | 0.9587 | 0.8514 | 0.9134 | 0.9355 | 0.8249 |
| Baseline + CheXpert | **0.9405** | **0.9689** | **0.9877** | **0.8624** | **0.9201** | **0.9542** | **0.8716** |
| Baseline + RSNA | 0.9359 | 0.9592 | **0.9877** | 0.8514 | 0.9158 | 0.9503 | 0.8653 |
| Baseline + NIH + CheXpert | 0.9333 | 0.9606 | 0.9835 | 0.8514 | 0.9154 | 0.9483 | 0.8594 |
| Baseline + NIH + RSNA | 0.9231 | 0.9642 | 0.9959 | 0.8041 | 0.8926 | 0.9415 | 0.8411 |
| Baseline + CheXpert + RSNA | 0.9359 | 0.9628 | 0.9835 | 0.8582 | 0.919 | 0.9501 | 0.8647 |
| Baseline + NIH + CheXpert + RSNA | 0.9154 | 0.9542 | 0.9794 | 0.8109 | 0.8944 | 0.935 | 0.8217 |
| Baseline + CheXpert + Twitter | 0.9103 | 0.9538 | 0.9629 | 0.8244 | 0.8997 | 0.9302 | 0.8088 |
| Baseline + CheXpert + Montreal | 0.9231 | 0.9595 | 0.9711 | 0.8446 | 0.9109 | 0.94 | 0.8365 |

Note that the baseline training data augmented with the weakly labeled CXR images from the CheXpert CXR collection demonstrated superior performance in all metrics compared to the non-augmented and other combinations of augmented training data. This underscores the fact that this augmentation approach resulted in a favorable increase in the training data size, encompassing a diversified distribution to learn and improve performance in the test data, compared to that with non-augmented training.

We also studied the effect of weakly labeled data augmentation with the test data from Twitter and Montreal COVID-19 CXR collections. The results are as shown in Table 6.

**Table 6.** Performance metrics achieved using combinations of the augmented training data toward classifying Twitter and Montreal COVID-19 CXR collections as belonging to the viral pneumonia category. Bold values indicate superior performance.

| Dataset | Accuracy |  |
| --- | --- | --- |
|  | Twitter-COVID-19 | Montreal-COVID-19 |
| Baseline | 0.2885 | 0.5028 |
| Data augmentation with weakly labeled images |  |  |
| Baseline + NIH | 0.1037 | 0.2625 |
| Baseline + CheXpert | 0.5555 | 0.6536 |
| Baseline + RSNA | 0.2296 | 0.4469 |
| Baseline + NIH + CheXpert | 0.1852 | 0.4078 |
| Baseline + NIH + RSNA | 0.1407 | 0.4413 |
| Baseline + CheXpert + RSNA | 0.2222 | 0.4357 |
| Baseline + NIH + CheXpert + RSNA | 0.1852 | 0.4413 |
| Baseline + CheXpert + Twitter | - | 0.7095 |
| Baseline + CheXpert + Montreal | 0.8889 | - |
| Baseline + Twitter | - | **0.9778** |
| Baseline + Montreal | **0.9926** | - |

The performance evaluation results demonstrate that the baseline training data augmented with the weakly labeled CXR images from the CheXpert collection initially improved performance with an accuracy of 0.5555 and 0.6536 as compared to the non-augmented baseline (0.2885 and 0.5028) in classifying Twitter and Montreal COVID-19 CXR test data, respectively, as belonging to the viral pneumonia category. The performance degradation with other combinations of weakly-labeled data augmentation underscores the fact that adding more data introduces noise into the training process; increasing the number of training samples do not always improve performance since these samples either do not contribute or adversely impact decision-making.

Modifying the distribution of the training data in a way to include only those samples could provide an effective solution to improve performance with the test data from a similar or different distribution as compared to the non-augmented training data. In this regard, we also augmented the baseline training data with the COVID-19 viral pneumonia CXRs from one of two different collections and evaluated the performance with the other. This is performed to evaluate if there is a performance improvement if the training data is modified to include only samples with a known, similar distribution. It is observed from Table 6 that augmenting the baseline training data with the Twitter COVID-19 CXR collection significantly improved performance in detecting COVID-19 CXRs in the Montreal collection as compared to the weakly-labeled augmentation using CheXpert CXRs and the non-augmented baseline. We observed similar improvements in performance with the Twitter COVID-19 CXRs when the baseline training data is augmented with the Montreal COVID-19 CXR collection for model training. Fig. 5 shows the confusion matrix obtained toward this study. This underscores the fact that augmenting the training data with COVID-19 CXRs, though not coming from the same collection, significantly improved performance with the test data from a different COVID-19 CXR collection, as compared to non-augmented baseline and weakly-labeled data augmentation with non-COVID-19 viral and bacterial pneumonia CXRs. The COVID-19 viral pneumonia has a distinct pattern, compared to non-COVID-19 viral and other pneumonia. For this reason, irrespective of the collection the CXRs come from, augmenting the training data with samples from one COVID-19 CXR collection significantly improves performance with the other.

**Figure 5.**
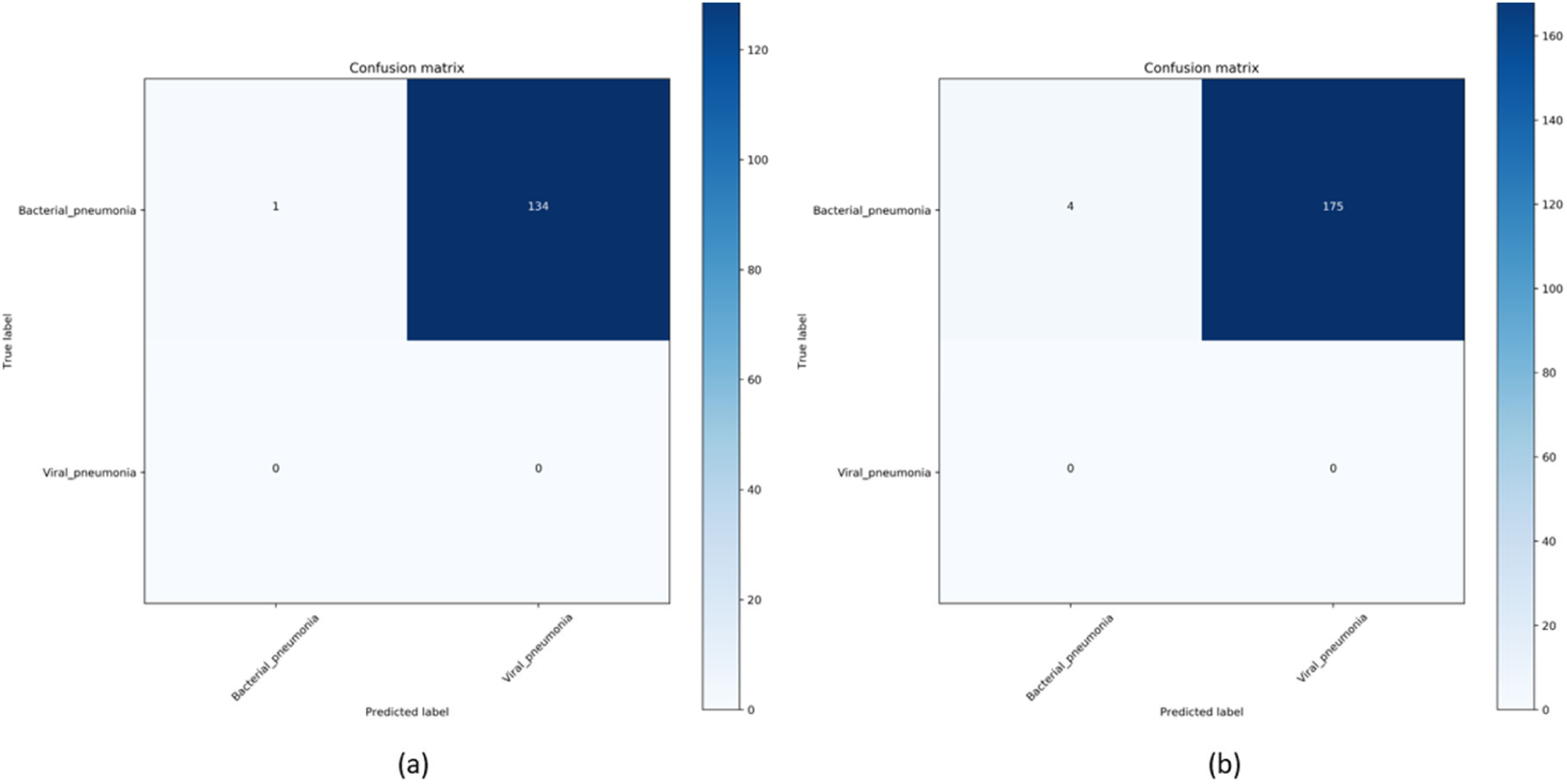
Confusion matrix obtained toward classifying (**a**) Twitter and (**b**) Montreal COVID-19 CXR collections as showing viral pneumonia using the VGG-16 model trained on the baseline augmented with Montreal COVID-19 and Twitter COVID-19 CXR collections, respectively.

Fig.6 shows the learned behavior of the VGG-16 model trained on the baseline data augmented with Montreal COVID-19 and Twitter COVID-19 CXR collections individually to predict on a test sample with GT annotations from Montreal COVID-19 and Twitter COVID-19 CXR collections, respectively.

**Figure 6.**
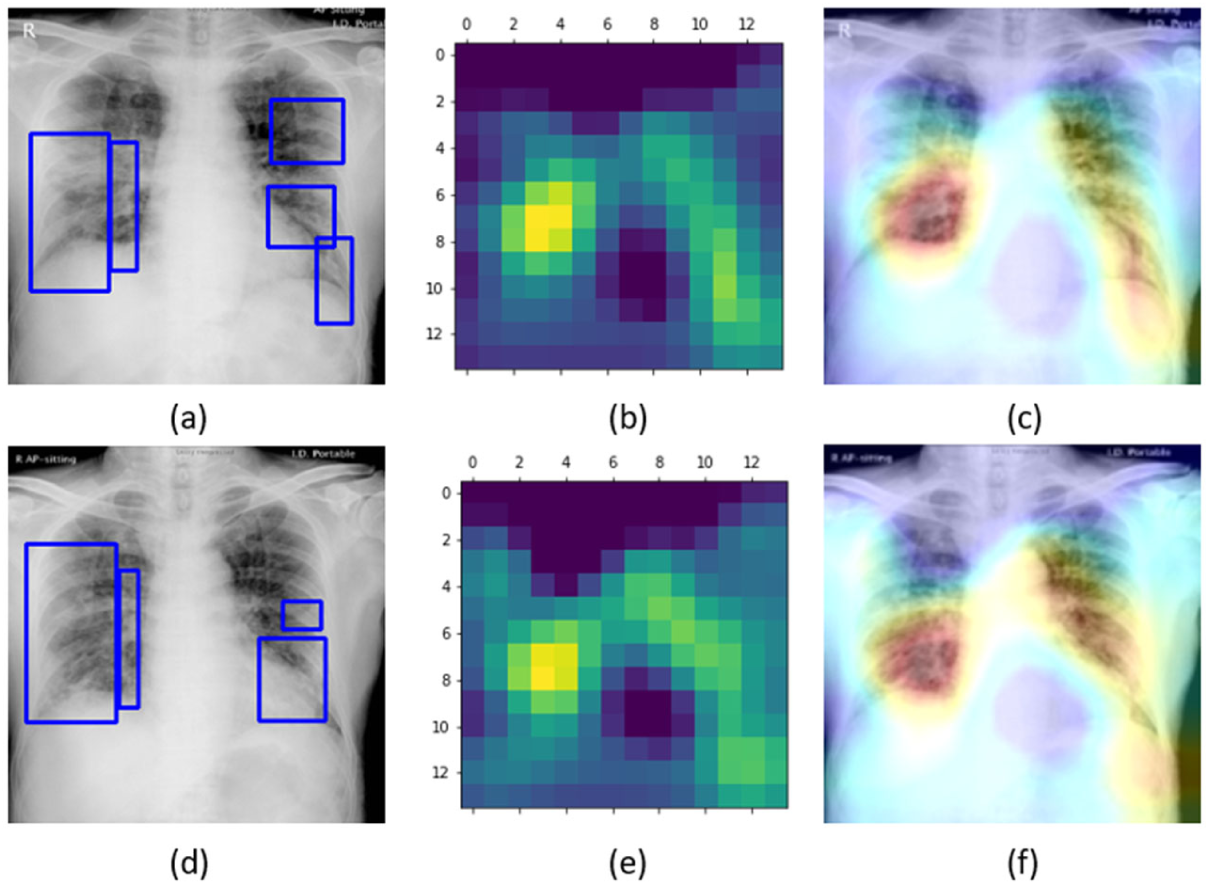
Original CXRs, heat maps, and salient ROI visualization: (**a**), (**b**), and (**c**) shows a COVID-19 viral pneumonia test CXR from Montreal collection with GT annotations, the corresponding heat map, and Grad-CAM visualization, (**d**), (**e**), and (**f**) shows a COVID-19 viral pneumonia test CXR from the Twitter collection with GT annotations, the heat map, and its associated class activation maps.

Unlike the degraded performance of the model trained on non-augmented data that failed to localize salient ROI in a test CXR showing COVID-19 viral pneumonia, as observed from Fig. 4, the model trained on the augmented baseline with COVID-19 CXRs from one collection delivered superior localization performance with the test CXR samples from the other collection. Fig. 6a shows the learned interpretation of these trained models in the form of heat maps and class activation maps. It is observed that the models are correctly focusing on the salient ROI, matching with the GT annotations that help to categorize them as showing COVID-19 viral pneumonia. This leads to the inference that the model has effectively learned the diversified feature space augmented with class-specific (COVID-19 viral pneumonia) data that has a distinct pattern compared to non-COVID-19 viral and bacterial pneumonia to effectively localize the salient ROI involved in decision-making.

## 4. Conclusions and Future Work

Image Weakly labeled data augmentation helped to improve performance with the hold-out baseline test data because the CXRs with pneumonia-related opacities in CheXpert collection has a similar distribution to bacterial and non-COVID-19 viral pneumonia that helped to expand the training feature space by introducing a controlled variance to improve performance with the baseline test data. However, with COVID-19 CXRs, weakly-labeled data augmentation didn’t deliver superior performance since COVID-19 viral pneumonia has a distinct pattern as compared to non-COVID-19 viral and bacterial pneumonia.

In this study, we evaluated the effect of weakly-labeled data augmentation toward classifying the CXRs as showing COVID-19 viral pneumonia. In this regard, being a one-class problem, we have only false-negatives and no false positives. As future work, we aim to expand the analysis toward classifying non-COVID-19 and COVID-19 viral pneumonia and other multi-class problems, where we aim to perform multi-class ROC analysis and obtain an efficient operating point suiting model deployment. Considering limited data availability as with COVID-19 detection, we also aim to construct model ensembles to combine the predictions of models trained on various combinations of augmented training data to further improve COVID-19 detection performance.

## Data Availability

The study uses publicly available datasets, the detailed are mentioned in the manuscript.

## Author Contributions

Conceptualization, Sivaramakrishnan Rajaraman; Data curation, Sivaramakrishnan Rajaraman; Formal analysis, Sivaramakrishnan Rajaraman; Funding acquisition, Sameer Antani; Investigation, Sivaramakrishnan Rajaraman and Sameer Antani; Methodology, Sivaramakrishnan Rajaraman and Sameer Antani; Project administration, Sameer Antani; Resources, Sameer Antani; Software, Sivaramakrishnan Rajaraman; Supervision, Sivaramakrishnan Rajaraman and Sameer Antani; Visualization, Sivaramakrishnan Rajaraman; Writing – original draft, Sivaramakrishnan Rajaraman; Writing – review & editing, Sivaramakrishnan Rajaraman and Sameer Antani. All authors have read and agreed to the published version of the manuscript.

## Acknowledgments

We are grateful to Dr. Jenifer Siegelman of Takeda Pharmaceuticals for her radiological expertise in annotating a sample of COVID-19 test data and discussions related to the radiology of COVID-19.

## Footnotes

1 https://www.acr.org/Advocacy-and-Economics/ACR-Position-Statements/Recommendations-for-Chest-Radiography-and-CT-for-Suspected-COVID19-Infection

2 https://press.rsna.org/timssnet/media/pressreleases/14_pr_target.cfm?ID=2167

3 https://imagingcovid19ai.eu/

